# Sex-specificity of mortality risk factors among hospitalized COVID-19 patients in New York City: prospective cohort study

**DOI:** 10.1101/2020.07.29.20164640

**Authors:** Tomi Jun, Sharon Nirenberg, Patricia Kovatch, Kuan-lin Huang

## Abstract

**Objective:** To identify sex-specific effects of risk factors for in-hospital mortality among COVID-19 patients admitted to a hospital system in New York City.

**Design:** Prospective observational cohort study with in-hospital mortality as the primary outcome.

**Setting:** Five acute care hospitals within a single academic medical system in New York City.

**Participants:** 3,086 hospital inpatients with COVID-19 admitted on or before April 13, 2020 and followed through June 2, 2020. Follow-up till discharge or death was complete for 99.3% of the cohort.

**Results:** The majority of the cohort was male (59.6%). Men were younger (median 64 vs. 70, p<0.001) and less likely to have comorbidities such as hypertension (32.5% vs. 39.9%, p<0.001), diabetes (22.6% vs. 26%, p=0.03), and obesity (6.9% vs. 9.8%, p=0.004) compared to women. Women had lower median values of laboratory markers associated with inflammation compared to men: white blood cells (5.95 vs. 6.8 K/uL, p<0.001), procalcitonin (0.14 vs 0.21 ng/mL, p<0.001), lactate dehydrogenase (375 vs. 428 U/L, p<0.001), C-reactive protein (87.7 vs. 123.2 mg/L, p<0.001). Unadjusted mortality was similar between men and women (28.8% vs. 28.5%, p=0.84), but more men required intensive care than women (25.2% vs. 19%, p<0.001). Male sex was an independent risk factor for mortality (OR 1.26, 95% 1.04-1.51) after adjustment for demographics, comorbidities, and baseline hypoxia. There were significant interactions between sex and coronary artery disease (p=0.038), obesity (p=0.01), baseline hypoxia (p<0.001), ferritin (p=0.002), lactate dehydrogenase (p=0.003), and procalcitonin (p=0.03). Except for procalcitonin, which had the opposite association, each of these factors was associated with disproportionately higher mortality among women.

**Conclusions:** Male sex was an independent predictor of mortality, consistent with prior studies. Notably, there were significant sex-specific interactions which indicated a disproportionate increase in mortality among women with coronary artery disease, obesity, and hypoxia. These new findings highlight patient subgroups for further study and help explain the recognized sex differences in COVID-19 outcomes.

## INTRODUCTION

Reports of the COVID-19 pandemic from around the world have described more severe disease and worse outcomes among men.[1–6] Men with COVID-19 appear to preferentially require hospitalization and intensive care.[2,7] One large Italian case series reported that 82% of COVID-19 patients requiring intensive care were men.[8] Case fatality rates also appear to be higher among men; a nationwide analysis from China reported a case fatality rate of 2.8% for men, compared to 1.7% for women.[4] Male sex has been identified as a risk factor for mortality in several studies of hospitalized COVID-19 patients.[9–13]

These observations have been variously attributed to underlying comorbidities among men, hormonal factors, or immune differences between men and women.[14–16] Some comorbidities associated with worse COVID-19 outcomes, such as cardiovascular disease and diabetes, may be more common among men, though published studies have not provided sex-disaggregated data for their cohorts.[2,17,18] In addition, there are sex-specific differences in expression of the ACE2 and TMPRSS2 proteins, which facilitate the entry of the SARS-CoV-2 virion into cells.[19,20] An Italian study further observed that among prostate cancer patients, those on androgen deprivation therapy had better outcomes than those who were not, suggesting a sex hormonal contribution to COVID-19 mortality.[21]

Given the apparent difference in COVID-19 outcomes between the sexes, we reasoned that clinical and molecular risk factors for mortality may show sex-specific effects, which remain largely uncharacterized. Although multiple prior studies have included multivariable regression models adjusting for sex, they have not explored the possibility of interactions between sex and other predictors. Models without sex interactions assume that predictors have the same effect on men’s and women’s mortality risks. However, given differences in immunity and comorbidities between men and women,[22–24] comorbid conditions and inflammatory responses may have different, sex-specific effects on COVID-19-associated mortality. To fill this gap in the literature, we undertook a sex-specific analysis of 3,086 COVID-19 patients hospitalized in New York City.

## METHODS

### Study setting

Mount Sinai Health System is a large hospital system in the New York City area, comprising 8 hospitals and more than 410 ambulatory practice locations. Our analysis focused on patients who were admitted to five hospitals: The Mount Sinai Hospital (1,134 beds), Mount Sinai West (514 beds), and Mount Sinai Morningside (495 beds) in Manhattan; Mount Sinai Brooklyn (212 beds) in Brooklyn; and Mount Sinai Queens (235 beds) in Queens.

### Data sources

Data were derived from clinical records from Mount Sinai facilities using the Epic electronic health record (Epic Systems, Verona, WI). Data were directly extracted from Epic’s Clarity and Caboodle servers. In the setting of the COVID-19 pandemic, the Mount Sinai Data Warehouse (MSDW) developed and released a de-identified data set encompassing all COVID-19 related patient encounters within the Mount Sinai system, accompanied by selected demographics, comorbidities, vital signs, medications, and lab values. As part of de-identification, all patients over the age of 89 had their age set to 90. Updated versions of the dataset have been released on a weekly schedule. For this study, we identified qualifying patient encounters from the dataset version released on April 13^th^, 2020 and followed their mortality outcomes through June 2^nd^, 2020.

This study utilized de-identified data extracted from the electronic health record and as such was considered non-human subject research. Therefore, this study was exempted from the Mount Sinai IRB review and approval process.

### Patient population and definitions

The MSDW dataset captured patient encounters at a Mount Sinai facility with any of the following: a COVID-19 related encounter diagnosis, a COVID-19 related visit type, a SARS-CoV-2 lab order, a SARS-CoV-2 lab result, or a SARS-CoV-2 lab test result from the New York State Department of Health’s Wadsworth laboratory.

We limited our analysis to adult patients (at least 18 years old) who were admitted for COVID-19 through the emergency department. Race and ethnicity were self-reported. COVID-19 positivity was defined as a positive or presumptive positive result from a nucleic acid-based test to detect SARS-CoV-2 in nasopharyngeal or oropharyngeal swab specimens. Initial vital signs were the first vital signs documented for the encounter. Oxygen saturation was dichotomized at a cutoff of 92% as this is the typical clinical threshold for initiating supplemental oxygen. We defined initial labs as the first lab value within 24 hours of the start of the encounter.

### Logistic regression analysis

The primary outcome was death from any cause during admission. Univariable and multivariable logistic regression were used to identify factors associated with mortality. To identify sex-specific effects of risk factors, the analysis was stratified by sex, and interaction terms were tested between sex and the covariates in the multivariable model.

Predictors analyzed included demographic factors, comorbidity status, initial vital signs, baseline lab values, and treatment facility site (Manhattan vs. Brooklyn/Queens). There was minimal clustering of the outcome by treatment site (ICC (ρ) = 0.026). Covariates were chosen a priori based on prior reports and low missingness. We report the odds ratios derived from the coefficients of each model, along with the Wald-type confidence interval and p-values.

Missingness of baseline lab values varied across sites and by test type. To avoid confounding by indication (lab tests selectively obtained on patients with more severe disease), we limited analyses involving lab tests to those obtained at the largest hospital site (The Mount Sinai Hospital) and which had less than 15% missing values at that site. We made exceptions for tests of interleukin-1-beta, interleukin-6, interleukin-8, and tumor necrosis factor-alpha because these were obtained as part of a study which enrolled consecutive COVID-19 patients hospitalized between March 21 and April 28, 2020.[25] Given the reduced sample size, the multivariable model applied to lab values omitted race, heart failure, and atrial fibrillation. Lab values were standardized (scaled to a mean of 0 and standard deviation of 1) for the analysis cohort prior to stratification and regression analysis.

### Statistical analysis

Patient characteristics and baseline vitals and labs were described using medians and ranges for continuous variables, and proportions for categorical variables. Continuous variables were compared using the Wilcoxon rank-sum test, and categorical variables were compared using Fisher’s exact test. All statistical analyses and data visualizations were carried out using R 4.0.0 (The R foundation, Vienna, Austria), along with the tidyverse, ggpubr, forestplot, and Hmisc packages. Statistical significance was defined as p<0.05.

### Patient and public involvement

Patients and the public were not directly involved in the design or conduct of the study, the choice of outcome measures, recruitment to the study, or the plan for dissemination of results.

## RESULTS

### Study population

The study cohort consisted of 3,086 COVID-positive patients admitted through the emergency department on or before April 13, 2020, and followed through June 2, 2020; 3,063 (99.3%) of patients were either discharged or died during the follow-up period (**Supplemental Figure 1**).

The cohort was predominantly male (59.1%) and racially diverse (28.9% Hispanic, 26.7% Non-Hispanic black, 22.3% Non-Hispanic white, and 4.7% Asian) (**Table 1**). There were significant baseline differences between men and women in the cohort. Men were younger (median age 64 vs. 74 years, p<0.001) and more likely to have a history of smoking (16.7% vs. 8.3%, p<0.001) than women. However, women were more likely to have pre-existing medical comorbidities such as hypertension (39.9% vs 32.5%, p<0.001), diabetes (26.0% vs. 22.6%, p=0.03), COPD/asthma (11.3% vs. 6.7%, p<0.001) and obesity (9.8% vs. 6.9%, p=0.004). Unadjusted mortality was similar between men and women (28.8% vs. 28.5%, p=0.84), but more men required intensive care than women (25.2% vs. 19%, p<0.001).

**Table 1:** Baseline characteristics, by sex

| Characteristic | Male<br>(N=1825) | Female<br>(N=1261) | P-value | Overall<br>(N=3086) |
| --- | --- | --- | --- | --- |
| Age (yrs) | 64 (54 - 75) | 70 (59 - 81) | <0.001 | 66 (56 - 77) |
| Race/ethnicity |  |  |  |  |
| Asian | 92 (5%) | 52 (4.1%) | 0.259 | 144 (4.7%) |
| Hispanic | 519 (28.4%) | 373 (29.6%) | 0.57 | 892 (28.9%) |
| Non-Hispanic Black | 445 (24.4%) | 380 (30.1%) | <0.001 | 825 (26.7%) |
| Non-Hispanic White | 429 (23.5%) | 260 (20.6%) | 0.0522 | 689 (22.3%) |
| Other | 290 (15.9%) | 168 (13.3%) | 0.0442 | 458 (14.8%) |
| **Treatment facility** |  |  |  |  |
| Mount Sinai Brooklyn (MSB) | 280 (15.3%) | 222 (17.6%) | 0.102 | 502 (16.3%) |
| Mount Sinai Queens (MSQ) | 420 (23%) | 223 (17.7%) | <0.001 | 643 (20.8%) |
| Mount Sinai Morningside (MSSL) | 319 (17.5%) | 251 (19.9%) | 0.0896 | 570 (18.5%) |
| Mount Sinai West (MSW) | 189 (10.4%) | 154 (12.2%) | 0.116 | 343 (11.1%) |
| The Mount Sinai Hospital (MSH) | 617 (33.8%) | 411 (32.6%) | 0.485 | 1028 (33.3%) |
| **Smoking status\*** |  |  |  |  |
| Current | 86 (4.7%) | 27 (2.1%) | <0.001 | 113 (3.7%) |
| Former | 429 (23.5%) | 229 (18.2%) | <0.001 | 658 (21.3%) |
| Never | 856 (46.9%) | 773 (61.3%) | <0.001 | 1629 (52.8%) |
| **Hypertension** | 593 (32.5%) | 503 (39.9%) | <0.001 | 1096 (35.5%) |
| **Diabetes** | 413 (22.6%) | 328 (26%) | 0.0321 | 741 (24%) |
| **Coronary artery disease** | 236 (12.9%) | 159 (12.6%) | 0.827 | 395 (12.8%) |
| **Heart failure** | 128 (7%) | 90 (7.1%) | 0.943 | 218 (7.1%) |
| **Atrial fibrillation** | 125 (6.8%) | 76 (6%) | 0.374 | 201 (6.5%) |
| **Chronic kidney disease** | 219 (12%) | 149 (11.8%) | 0.91 | 368 (11.9%) |
| **COPD/asthma** | 122 (6.7%) | 143 (11.3%) | <0.001 | 265 (8.6%) |
| **Obesity** | 126 (6.9%) | 124 (9.8%) | 0.00386 | 250 (8.1%) |
| **Cancer** | 125 (6.8%) | 80 (6.3%) | 0.607 | 205 (6.6%) |
| **Chronic liver disease** | 55 (3%) | 28 (2.2%) | 0.213 | 83 (2.7%) |
| **Obstructive sleep apnea** | 40 (2.2%) | 25 (2%) | 0.799 | 65 (2.1%) |
| **HIV** | 42 (2.3%) | 14 (1.1%) | 0.0189 | 56 (1.8%) |
| **Initial vitals** |  |  |  |  |
| Temperature (°F) | 99 (98.2 - 100.4) | 98.8 (98.2 - 100.1) | 0.00933 | 98.9 (98.2 - 100.2) |
| Heart rate (bpm) | 98 (85 - 111) | 95 (83 - 108) | <0.001 | 97 (84 - 110) |
| Systolic blood pressure (mmHg) | 130 (117 - 147) | 128 (114 - 144) | 0.00115 | 129 (115 - 146) |
| Respiratory rate (bpm) | 20 (18 - 22) | 20 (18 - 22) | 0.037 | 20 (18 - 22) |
| Oxygen saturation (%) | 94 (91 - 97) | 95 (92 - 97) | <0.001 | 95 (91 - 97) |

Values represent count (%) or median (IQR) for categorical and continuous variables, respectively
\* 22% missing values

### Sex-specific mortality risk factors

Male sex was not associated with mortality in univariable analysis (OR 1.02, 95% CI 0.868-1.19) (**Supplemental Table 1**). However, male sex was an independent predictor of mortality (OR 1.26, 95% CI 1.04-1.51) in a multivariable model controlling for age, race, comorbidities, treatment location, and initial oxygen saturation (**Table 2**).

**Table 2:**
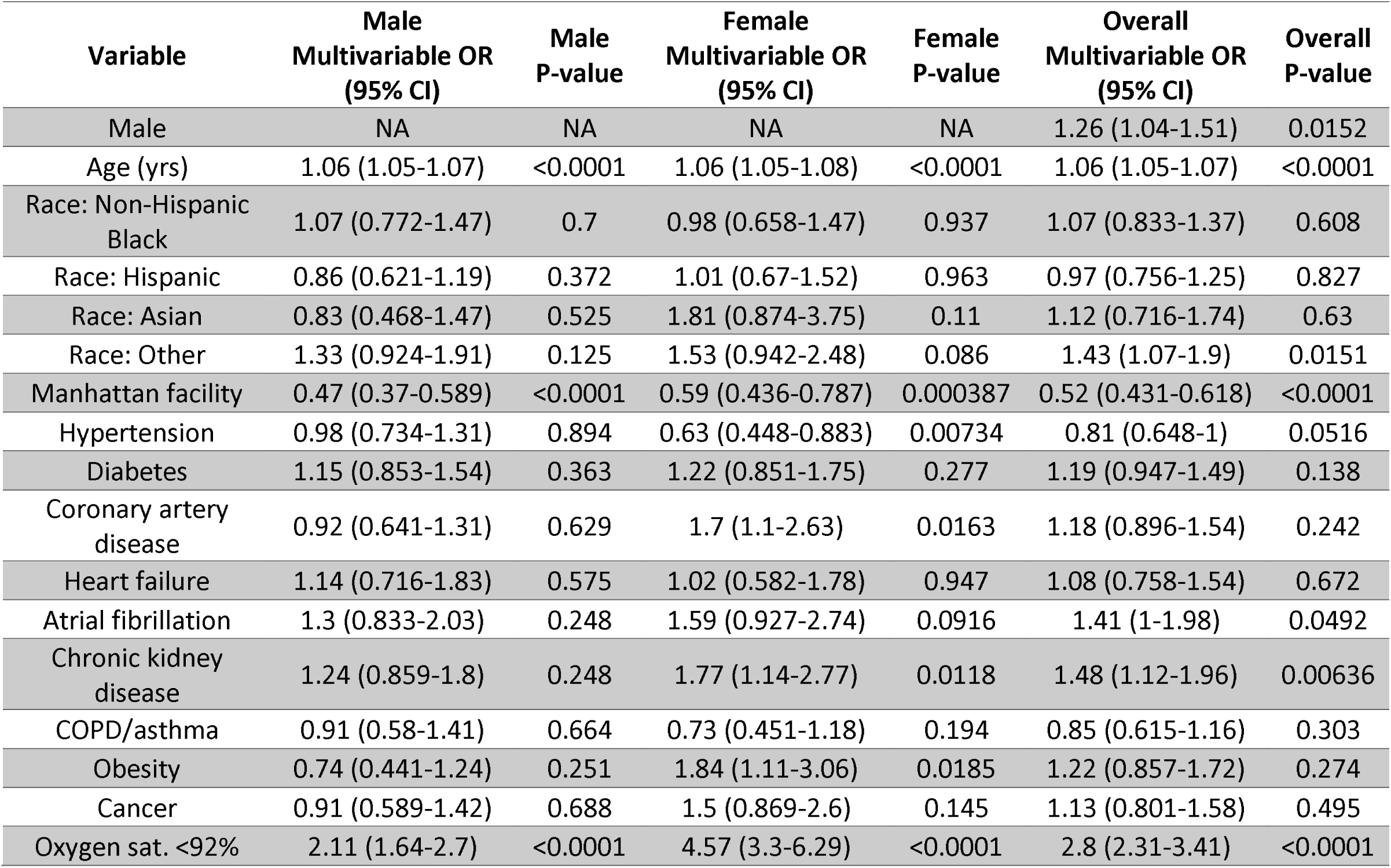
Multivariable regression predicting inpatient mortality, by sex

| Variable | Male<br>Multivariable OR<br>(95% CI) | Male<br>P-value | Female<br>Multivariable OR<br>(95% CI) | Female<br>P-value | Overall<br>Multivariable OR<br>(95% CI) | Overall<br>P-value |
| --- | --- | --- | --- | --- | --- | --- |
| Male | NA | NA | NA | NA | 1.26 (1.04-1.51) | 0.0152 |
| Age (yrs) | 1.06 (1.05-1.07) | <0.0001 | 1.06 (1.05-1.08) | <0.0001 | 1.06 (1.05-1.07) | <0.0001 |
| Race: Non-Hispanic<br>Black | 1.07 (0.772-1.47) | 0.7 | 0.98 (0.658-1.47) | 0.937 | 1.07 (0.833-1.37) | 0.608 |
| Race: Hispanic | 0.86 (0.621-1.19) | 0.372 | 1.01 (0.67-1.52) | 0.963 | 0.97 (0.756-1.25) | 0.827 |
| Race: Asian | 0.83 (0.468-1.47) | 0.525 | 1.81 (0.874-3.75) | 0.11 | 1.12 (0.716-1.74) | 0.63 |
| Race: Other | 1.33 (0.924-1.91) | 0.125 | 1.53 (0.942-2.48) | 0.086 | 1.43 (1.07-1.9) | 0.0151 |
| Manhattan facility | 0.47 (0.37-0.589) | <0.0001 | 0.59 (0.436-0.787) | 0.000387 | 0.52 (0.431-0.618) | <0.0001 |
| Hypertension | 0.98 (0.734-1.31) | 0.894 | 0.63 (0.448-0.883) | 0.00734 | 0.81 (0.648-1) | 0.0516 |
| Diabetes | 1.15 (0.853-1.54) | 0.363 | 1.22 (0.851-1.75) | 0.277 | 1.19 (0.947-1.49) | 0.138 |
| Coronary artery<br>disease | 0.92 (0.641-1.31) | 0.629 | 1.7 (1.1-2.63) | 0.0163 | 1.18 (0.896-1.54) | 0.242 |
| Heart failure | 1.14 (0.716-1.83) | 0.575 | 1.02 (0.582-1.78) | 0.947 | 1.08 (0.758-1.54) | 0.672 |
| Atrial fibrillation | 1.3 (0.833-2.03) | 0.248 | 1.59 (0.927-2.74) | 0.0916 | 1.41 (1-1.98) | 0.0492 |
| Chronic kidney<br>disease | 1.24 (0.859-1.8) | 0.248 | 1.77 (1.14-2.77) | 0.0118 | 1.48 (1.12-1.96) | 0.00636 |
| COPD/asthma | 0.91 (0.58-1.41) | 0.664 | 0.73 (0.451-1.18) | 0.194 | 0.85 (0.615-1.16) | 0.303 |
| Obesity | 0.74 (0.441-1.24) | 0.251 | 1.84 (1.11-3.06) | 0.0185 | 1.22 (0.857-1.72) | 0.274 |
| Cancer | 0.91 (0.589-1.42) | 0.688 | 1.5 (0.869-2.6) | 0.145 | 1.13 (0.801-1.58) | 0.495 |
| Oxygen sat. <92% | 2.11 (1.64-2.7) | <0.0001 | 4.57 (3.3-6.29) | <0.0001 | 2.8 (2.31-3.41) | <0.0001 |

We tested interactions between sex and each of the other predictors in the multivariable model and found significant interactions between sex and coronary artery disease (CAD, p=0.038), obesity (p=0.01), and baseline hypoxia (oxygen saturation <92%, p<0.001) (**Figure 1**). In the overall model, baseline hypoxia was independently associated with mortality (OR 2.8, 95% CI 2.31-3.41), but CAD (OR 1.18, 95% CI 0.896-1.54) and obesity (OR 1.22, 95% CI 0.857-1.72) were not. In the sex-stratified models, each of these factors was associated with increased mortality in women to a greater degree than in men (**Figure 1**).

**Figure 1:**
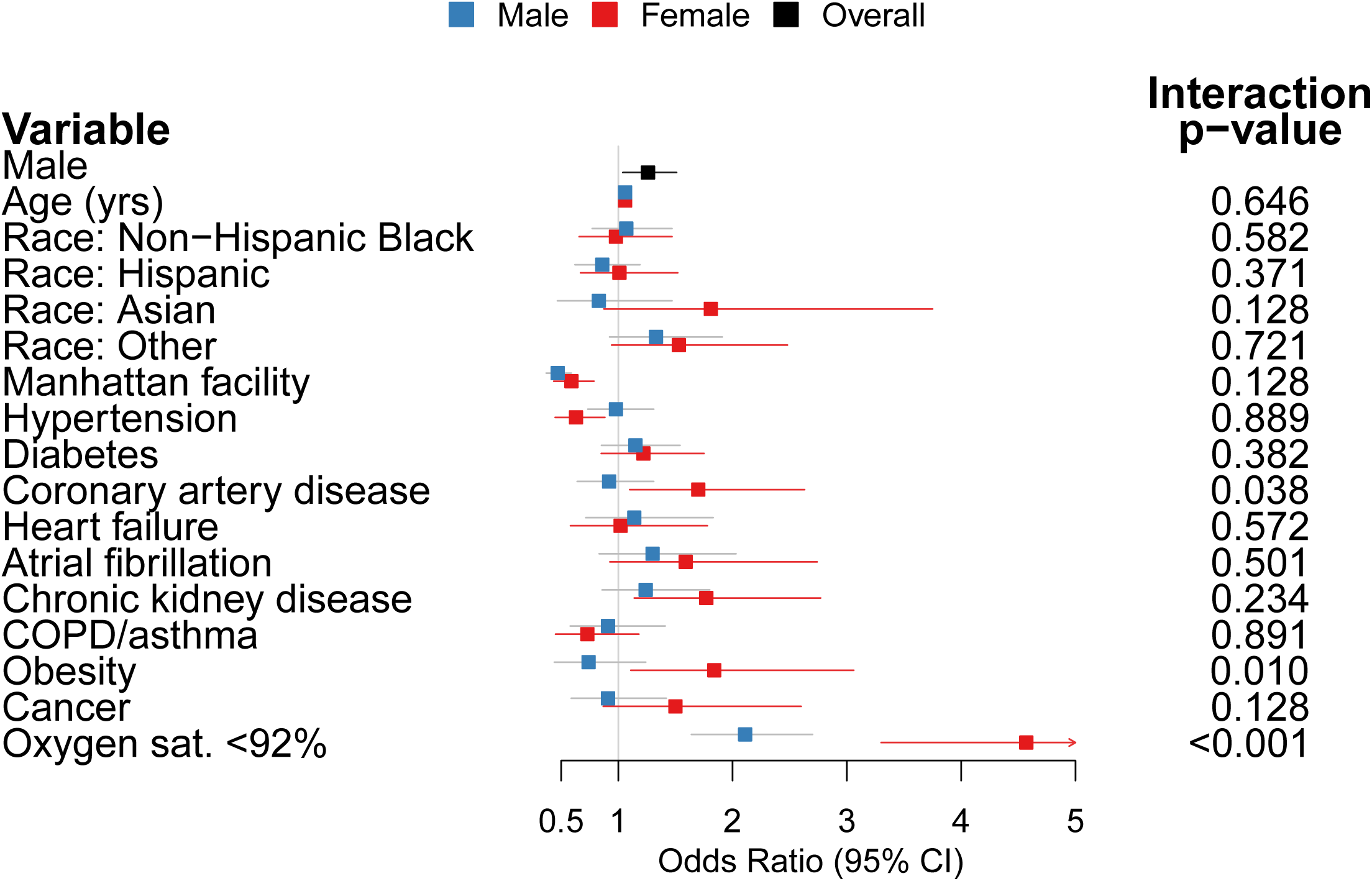
Forest plot of multivariable logistic regression model predicting in-hospital mortality, stratified by sex

CAD and obesity were not independent mortality risk factors among men (CAD OR 0.92, 95% CI 0.641-1.31; Obesity OR 0.74, 95% CI 0.441-1.24), but they were independent risk factors among women (CAD OR 1.7, 95% CI 1.1-2.63; Obesity OR 1.84, 95% CI 1.11-3.06). Baseline hypoxia was associated with increased mortality in both men (OR 2.11, 95% CI 1.64-2.7) and women (OR 4.57, 95% CI 3.3-6.29), but had a greater impact on women’s mortality risk. Considered another way, male sex (compared to female sex) was associated with increased mortality among normoxic patients (OR 1.55, 95% CI 1.24-1.93), but not among hypoxic patients (OR 0.76, 95% CI 0.54-1.08).

### Sex differences in baseline lab values and associations with mortality

We conducted an analysis of baseline lab values among the subset of patients seen at the largest site (The Mount Sinai Hospital, N=1,028), focusing on lab values that were collected routinely at baseline. We also analyzed baseline cytokine levels, which were collected on a subset of consecutive patients in this population (N=610).

At baseline, women exhibited lower median values of several laboratory markers associated with inflammation compared to men: white blood cells (WBC; 5.95 vs. 6.8 K/uL, p<0.001), procalcitonin (0.14 vs 0.21 ng/mL, p<0.001), lactate dehydrogenase (LDH; 375 vs. 428 U/L, p<0.001), C-reactive protein (CRP; 87.7 vs. 123.2 mg/L, p<0.001) (**Table 3, Supplemental Figure 2**). Baseline ferritin was lower in women than in men in absolute terms (459.5 vs. 958 ng/mL, p<0.001), but relative to the upper limit of normal (ULN) for each sex, women had more markedly elevated ferritin levels (3.06 vs. 2.40 x ULN, p=0.005).

**Table 3:** Baseline laboratory values among patients at The Mount Sinai Hospital, by sex

| Laboratory test | N | Male | Female | P-value | Overall |
| --- | --- | --- | --- | --- | --- |
| White blood cells, $10^3/\mu\text{L}$ | 1027 | 6.8 (5.2 - 9.9) | 5.95 (4.725 - 8.7) | $<0.001$ | 6.5 (5 - 9.4) |
| Hemoglobin, g/dL | 1028 | 13.4 (12.1 - 14.6) | 12.3 (11 - 13.5) | $<0.001$ | 13.1 (11.6 - 14.2) |
| Platelets, $10^3/\mu\text{L}$ | 1023 | 204 (157.5 - 263.5) | 207 (166.75 - 267) | 0.307 | 205 (160 - 264.5) |
| Sodium, mmol/L | 1027 | 136 (133 - 139) | 138 (135 - 141) | $<0.001$ | 137 (134 - 140) |
| Potassium, mmol/L | 984 | 4.2 (3.8 - 4.7) | 4.1 (3.7 - 4.6) | 0.00697 | 4.2 (3.8 - 4.6) |
| Chloride, mmol/L | 1027 | 101 (97 - 104) | 103 (99 - 106) | $<0.001$ | 101 (98 - 105) |
| Blood urea nitrogen, mg/dL | 1027 | 18 (12 - 30) | 17 (10 - 32) | 0.019 | 17 (11 - 31) |
| Creatinine, mg/dL | 1028 | 1.02 (0.81 - 1.51) | 0.83 (0.63 - 1.435) | $<0.001$ | 0.95 (0.73 - 1.5) |
| Aspartate aminotransferase, U/L | 955 | 46 (31 - 71) | 37.5 (28 - 57.25) | $<0.001$ | 41 (29.5 - 64) |
| Alanine aminotransferase, U/L | 999 | 34 (20 - 60) | 25 (17 - 37) | $<0.001$ | 30 (19 - 51) |
| Total bilirubin, mg/dL | 1007 | 0.6 (0.5 - 0.9) | 0.5 (0.4 - 0.7) | $<0.001$ | 0.6 (0.4 - 0.8) |
| Albumin, g/dL | 1010 | 3.2 (2.8 - 3.5) | 3.3 (2.9 - 3.6) | 0.0908 | 3.2 (2.9 - 3.5) |
| D-dimer, $\mu\text{g/mL FEU}$ | 792 | 1.32 (0.74 - 2.365) | 1.19 (0.7 - 2.21) | 0.325 | 1.29 (0.71 - 2.3125) |
| Ferritin, ng/mL | 943 | 958 (462 - 2108.5) | 459.5 (222 - 892.75) | $<0.001$ | 700 (339.5 - 1684) |
| Procalcitonin, ng/mL | 957 | 0.21 (0.1 - 0.58) | 0.14 (0.06 - 0.5275) | $<0.001$ | 0.18 (0.08 - 0.55) |
| Lactate dehydrogenase, U/L | 892 | 428 (326 - 560) | 375.5 (303.75 - 497.5) | $<0.001$ | 402 (313 - 533.25) |
| C-reactive protein, mg/L | 944 | 123.2 (65.3 - 196.7) | 87.7 (45.25 - 148.15) | $<0.001$ | 107.95 (55.625 - 179.975) |
| Interleukin-1 beta, pg/mL | 437 | 0.5 (0.4 - 0.9) | 0.5 (0.4 - 0.8) | 0.672 | 0.5 (0.4 - 0.8) |
| Interleukin-6, pg/mL | 610 | 85.8 (47 - 144.3) | 59.9 (36.1 - 122) | 0.00148 | 76.05 (40.8 - 138) |
| Interleukin-8, pg/mL | 543 | 41 (25.025 - 63.925) | 41.6 (27.7 - 64.9) | 0.529 | 41.5 (25.75 - 64.05) |
| Tumor necrosis factor-alpha, pg/mL | 544 | 22.7 (16.85 - 33.05) | 23.3 (16.9 - 33.9) | 0.874 | 22.8 (16.875 - 33.225) |

Baseline interleukin-8 (IL-8; median 41.5 pg/mL; reference range 0-5 pg/mL) and interleukin-6 (IL-6; median 76.1 pg/mL; reference range 0-5 pg/mL) levels were elevated among all patients, while tumor necrosis factor alpha (TNFa; median 22.8 pg/mL; reference range: 0-22 pg/mL) and interleukin-1 beta (IL-1B; median 0.5 pg/mL; reference range 0-5 pg/mL) levels were not. Only IL-6 exhibited a sex difference, with women having lower median baseline levels than men (59.9 vs. 85.8 pg/mL, p=0.001) (**Supplemental Figure 2H**).

We assessed the association of selected inflammatory markers and cytokines with in-hospital mortality using multivariable models adjusting for age, sex, comorbidities, and initial oxygen saturation (**Supplemental Table 2**). Higher baseline values of white blood cells (p=0.002), C-reactive protein (p<0.001), lactate dehydrogenase (p<0.001), procalcitonin (p=0.01), d-dimer (p=0.02), IL-6 (p<0.001), and IL-8 (p=0.002) were independently associated with increased mortality. Albumin, a negative acute phase reactant, was negatively correlated with mortality (p=0.001).

### Interactions between inflammatory markers and sex or hypoxia

Given the interaction between sex and baseline hypoxia, we also tested interactions between these two variables and the inflammatory markers. There were significant interactions between sex and ferritin (p=0.002), LDH (p=0.003), and procalcitonin (p=0.03) (**Figure 2A**). In a sex-stratified analysis of standardized lab values, ferritin was not associated with mortality among men (OR 0.947, 95% CI 0.776-1.16), but was in women (OR 1.66, 95% CI 1.2-2.29). Procalcitonin was associated with mortality in men (OR 2.34, 95% CI 1.19-4.61), but not in women (or 1.05, 95% CI 0.85-1.29). LDH was associated with mortality in both men (OR 1.26, 95% CI 1.01-1.57) and women (OR 2.50, 95% CI 1.66-3.79), but was a stronger predictor among women (**Supplemental Table 2**). Because standardized values were used in these regressions, the odds ratios should be interpreted as the change in odds associated with a one-standard-deviation increase in the value of the test.

**Figure 2:**
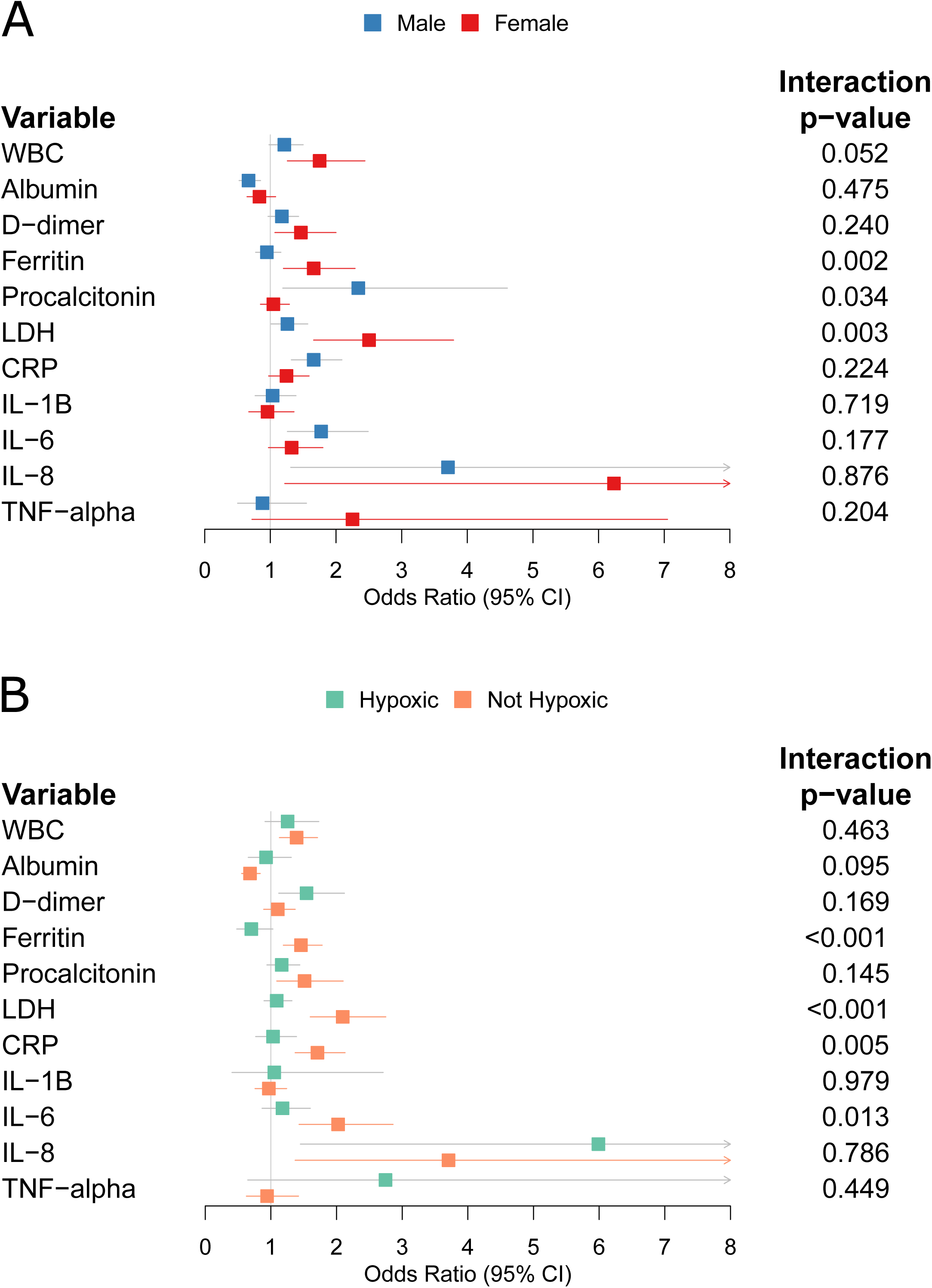
Forest plots of multivariable logistic regression models predicting in-hospital mortality, using baseline laboratory values as predictors, stratified by (A) sex, and (B) baseline hypoxia

There were significant interactions between baseline hypoxia and LDH (p<0.001), ferritin (p<0.001), CRP (p=0.005), and IL-6 (p=0.01) (**Figure 2B**). All four markers were associated with increased mortality among patients who were not hypoxic at baseline (LDH OR 2.09, 95% CI 1.6-2.75; Ferritin OR 1.46, 95% CI 1.19-1.78; CRP OR 1.71, 95% CI 1.37-2.13; IL6 OR 2.02, 95% CI 1.43-2.86). However, the markers were not associated with mortality among those who were hypoxic at baseline (LDH OR 1.09, 95% CI 0.896-1.32; Ferritin OR 0.70, 95% CI 0.48-1.03; CRP OR 1.03, 95% CI 0.768-1.39; IL6 OR 1.18, 95% CI 0.866-1.6). (**Supplemental Table 3**).

## DISCUSSION

As the COVID-19 pandemic has progressed, patient cohort studies have consistently identified a difference in mortality outcomes between men and women infected with SARS-CoV-2. However, it remains unclear what underlies the sex difference.

Like other studies from the US,[2,5] the UK,[6] Italy,[1] and China,[9] we found that male sex was an independent risk factor for mortality. However, prior studies have not provided sex-disaggregated data. When stratifying by sex, we found that raw mortality rates were similar between men and women (28.8% vs. 28.5%), even though men in this cohort were significantly younger and had fewer comorbidities than women (**Table 1**). This observation further illustrates the disparity in COVID-19 outcomes between men and women.

By testing interaction terms and using sex-stratified models, we identified obesity and coronary artery disease (CAD) as comorbidities that disproportionately increased women’s mortality risk. Both obesity and CAD have been identified in other studies as mortality risk factors.[2,3,6] However, this is the first study to report these sex-specific associations. Obese women and those with a history of CAD may represent higher risk subgroups relative to other women.

Additionally, baseline hypoxia (oxygen saturation <92%) interacted significantly with sex. Men were more likely to present with hypoxia (27.0% vs. 22.4%, p=0.004), but, compared to individuals of the same sex with normal oxygen saturation, mortality was 4.5 times higher among hypoxic women, and 2.1 times higher among hypoxic men. Due to this interaction, men and women presenting with hypoxia had a similar mortality risk; the protective association of female sex was abolished in the presence of hypoxia. This finding adds nuance to the use of hypoxia severity to categorize COVID-19 severity among patients in therapeutic trials: women without hypoxia had lower mortality risk than men without hypoxia, but men and women with hypoxia had similar risks, all else being equal. Thus, the risk-benefit ratio of an early-intervention agent administered to non-hypoxic patients may differ for men and women, whereas the risk-benefit ratio would not differ by sex for a therapy administered to hypoxic patients.

To further delineate the interactions between sex, disease severity, and inflammation, we analyzed baseline inflammatory markers in the cohort. Women had lower levels of inflammatory markers and cytokines than men at baseline. Overall, inflammation was associated with mortality, as evidenced by the independent associations of inflammatory markers such as WBCs, CRP, LDH, procalcitonin, d-dimer, IL-6 and IL-8 with mortality. Interestingly, four of these markers –LDH, ferritin, CRP, and IL-6 – had significant interactions with hypoxia. Patients without hypoxia, but with elevated levels of these markers, had disproportionately higher mortality. This increased-risk subgroup warrants further validation and evaluation.

The strengths of our study include its size and the diversity of the patient population, along with complete follow-up of 99.3% of patients in the cohort. Although larger, national-level patient cohorts have been published,[6,26] this Mount Sinai Health System cohort is on par with other health-system and hospital-network-based cohorts from the US and Italy.[1–3,5,7] This cohort empowers us to explicitly address sex differences in mortality risk factors using a rigorous combination of stratification and interaction models. To our knowledge, we are the first to report sex-stratified data of risk factors, inflammatory markers, and their association with mortality.

This study has several limitations. Our analyses are hypothesis-generating and require confirmation in other large cohorts as well as causal studies to establish mechanisms. Comorbidity data was automatically extracted from the electronic medical record and may be incomplete or inaccurate. This cohort reflects patients seen in the Spring of 2020 leading up to and during the initial surge of cases in New York City. Demographics and outcomes of COVID-19 patients may shift over the course of the pandemic.

In conclusion, the findings of this large study highlight novel interactions between sex, comorbidities, hypoxia, and markers of inflammation. These interactions nominate patient subgroups for further study and provide insights that may explain the recognized sex differences in outcomes of this disease.

## Data Availability

Patient level data is not available for public dissemination.

## ACKNOWLEDGEMENTS

We dedicate this work to the frontline health workers and staff of the Mount Sinai Healthcare System. This work was supported in part through the Mount Sinai Data Warehouse (MSDW) resources and staff expertise provided by Scientific Computing at the Icahn School of Medicine at Mount Sinai.

## AUTHOR CONTRIBUTIONS

TJ and KH conceived the idea and designed the study. SN and PK collected the data. TJ and KH conducted the statistical analysis. All authors interpreted the results. TJ drafted the initial manuscript and KH edited the manuscript. All authors revised and approved the manuscript.

## Supplemental Materials

Supplemental Table 1: Univariable logistic regression results with in-hospital mortality as the outcome

Supplemental Table 2: Multivariable logistic regression results for inflammatory markers as predictors of in-hospital mortality, adjusting for age, comorbidities, and initial oxygen saturation, stratified by sex. The unstratified model includes sex. Interaction P-values indicate the significance of the interaction term between sex and the lab value in a multivariable model incorporating the interaction. Lab values were standardized prior to stratification and regression; odds ratios represent the change in odds associated with a one-standard-deviation increase in the lab value.

Supplemental Table 3: Multivariable logistic regression results for inflammatory markers as predictors of in-hospital mortality, adjusting for age, sex, comorbidities, stratified by initial oxygen saturation. The unstratified model includes initial oxygen saturation. Interaction P-values indicate the significance of the interaction term between sex and the lab value in a multivariable model incorporating the interaction. Lab values were standardized prior to stratification and regression; odds ratios represent the change in odds associated with a one-standard-deviation increase in the lab value.

Supplemental Figure 1: Flow diagram of included patients

*Supplemental Figure 2: Baseline lab values among patients admitted to The Mount Sinai Hospital, by sex. (A) White blood cells. (B) Procalcitonin. (C) lactate dehydrogenase. (D) C-reactive protein. (E) Ferritin, absolute values. (F) Ferritin, relative to upper limit of normal. (G) Interleukin-8. (H) Interleukin-6. (I) Tumor necrosis factor alpha. (J) Interleukin-1 beta. Dotted lines represent reference ranges. Female-specific reference ranges are in red while male-specific reference ranges are in blue. Significance levels are indicated by ns: p > 0*.*05, **: p ≤ 0*.*01, ***: p ≤ 0*.*001; ****: p ≤ 0*.*0001*.

